# Recovery and discharge time between remimazolam tosylate and propofol in hysteroscopy anesthesia: a randomized controlled study

**DOI:** 10.1101/2024.04.10.24305650

**Authors:** Xiangyi Lin, Shengjie Chen, Huiyan Liang

**Author notes:** Corresponding Author: Shengjie Chen ^1^, Department of Anesthesiology, Maternal and Child Health Hospital of Shunde Foshan, Shunde Women and Children’s Hospital of Guangdong Medical University, NO.3 Baojian Road Street, Foshan, Guangdong, 528300, China.

## Abstract

**Background:** Hysteroscopy is considered the gold standard for the evaluation and treatment of uterine and endometrial lesions, but the operation is accompanied by severe pain, so it needs to be performed under anesthesia. Currently, a new sedative drug, remimazolam tosilate, can be used in digestive endoscopy and bronchoscopy. We will study its effect on hysteroscopy and its impact on patient recovery time and hospital discharge time.

**Methods:** Ninety-five patients undergoing hysteroscopy were randomly divided into two groups and received an initial dose of 0.2 mg/kg remimazolam tosilate (R group) and an initial dose of 2 mg/kg propofol (P group) as anesthetics during the examination. The primary end point was anesthesia recovery time (the time required for the patient’s MOAA/S score to reach 5 from the cessation of drug use) and hospital discharge time (the time required for the patient’s PADS score to reach 9 from the cessation of drug use). Secondary endpoints were first-dose sedation success rate, incidence of hypotension, and injection pain.

**Results:** Age(*P =* 0.825), height(*P =* 0.174), weight(*P =* 0.667) and ASA grade (*P =* 0.972) were not significantly different between the two groups. The recovery time in the R group was significantly shorter than that in the P group (*P =* 0.019), and the data kurtosis in the R group was higher than that in the P group (1.19 vs −0.56), but there was no significant difference in the time to hospital discharge (*P =* 0.696). There was no significant difference in the success rate of the first dose of sedation between the two groups (*P =* 0.362), but the incidence of hypotension during anesthesia (*P =* 0.001) and the incidence of injection pain (*P =* 0.001) in the R group was significantly lower than those in P group.

**Conclusions:** Remimazolam tosilate is suitable for hysteroscopy anesthesia and can reduce intraoperative hypotension and injection pain during the examination, a more optimal dosage or target-controlled infusion model still needs to be further explored.

## Introduction

Hysteroscopy is considered to be the gold standard for evaluating and managing uterine and endometrial lesions (van Hanegem et al. 2016; Wang et al. 2021), but cervical dilation and endometrial curettage are accompanied by severe pain(Riemma et al. 2020). Therefore, most hysteroscopies are performed under anesthesia. The most commonly used sedative drug is propofol due to its fast effect(Yu et al. 2019), easy titration, and short half-life, and can also reduce postoperative nausea and vomiting(Schraag et al. 2018). But its large impact on hemodynamics, no specific antagonists, and accumulation after long-term infusion have been its significant adverse effects in clinical application. As a novel ultra-short-acting benzodiazepine sedative drug, remimazolam tosilate has been successfully used as a sedative in bronchoscopy(Jia et al. 2021) and gastrointestinal endoscopy(Rex et al. 2018). It is designed to be rapidly hydrolyzed in the body by ubiquitous tissue esterases to an inactive carboxylic acid metabolite(Kilpatrick et al. 2007). Compared with the existing benzodiazepines, it has a faster onset of action, a shorter and predictable duration of sedation, and outstanding advantages in short-term anesthesia and operation.

This study hopes to provide patients with more comfort and shorten treatment time by analyze and comparing the differences in recovery time, hospital discharge time and injection pain indicators between remimazolam tosilate and propofol in anesthesia for hysteroscopy.

## Materials & Methods

### Study Design and Participants

This study is a single-center, parallel randomized controlled study, which was approved by the Medical Research Ethics Committee of Shunde Women and Children’s Hospital of Guangdong Medical University (Maternal and Child Health Hospital of Shunde Foshan) (registration number:2020055) and registered on Chictr.org (Date of Registration:15/07/2021, Registration number:ChiCTR2100048738). The study protocol followed the CONSORT and related clinical guidelines. All patients participating in the study signed written informed consent of participation and publication.

Inclusion criteria for this study: 1. planning to undergo hysteroscopy without endotracheal intubation; 2. aged 18-50 years; 3. American Society of Anesthesiologists (ASA) Physical Status rating I-II. Exclusion criteria: 1. Underweight or overweight (BMI>=30 or <18.5); 2. Anemia (HGB≤90g/L); 3. Severe liver and kidney dysfunction; 4. History of sedative abuse and mental illness; 5. Pregnancy or Within 3 months of breastfeeding; 6. Those who intend to undergo spinal anesthesia during examination and operation and those who refuse to participate in this study.

### Randomization and Blinding

In this study, the block randomization method was adopted, and R software was used to generate a random sequence based on 4 patients as a block, and the allocation ratio is 1:1. The results were put into opaque envelopes. After the patients signed the informed consent, the researchers unsealed the corresponding envelopes, and allocated the patients. Enter the remimazolam group (R group) and propofol group (P). The study was blinded to the follow-up staff.

### Research intervention

All patients received intravenous anesthesia, and patient monitor (iPM8, Mindray Biomedical Electronics Co., Ltd., Shenzhen, China) was used to measure non-invasive blood pressure, SpO2, ECG and RESP during anesthesia and recovery. After monitoring and venous catheter placement, the initial dose of remimazolam tosilate (Jiangsu Hengrui Pharmaceutical Co., Ltd., China) in R group was 0.2 mg/kg, followed by infusion at a rate of 1 mg/kg*hr, and an additional 0.1 mg/kg remimazolam was given for body movement during the examination The rescue dose was given no more than once every 5 minutes; the initial dose of propofol (Zhejiang Nhwa Pharmaceutical Co., Ltd., China) in P group was 2 mg/kg, followed by infusion at a rate of 8 mg/kg*hr, and an additional 1 mg/kg propofol was given to those who experienced body movement during the examination. The rescue dose was given no more than once every 5 minutes. At the end of the examination, flurbiprofen axetil (Grand Life Sciences(Wuhan) Co., Ltd., China) 50 mg was administered intravenously for postoperative analgesia.

### Sample size and statistical analysis

In a pilot study of hysteroscopy using propofol and remimazolam(Zhang et al. 2021), patients spent 5.44 minutes (remimazolam group) and 6.3 minutes (propofol group) in the PACU, with a standard deviation of 1.45 . A sample size of 45 participants per group was calculated with a significance level of 0.05 (a = 0.05), a strength of 80% (b = 0.20), and a dropout rate of 10%.

Statistical analysis was performed using SPSS Statistics 25.0 (SPSS Inc., Chicago, IL). Normality test in SPSS statistics software was used for data analysis to determine whether the data were in accordance with a normal distribution. Normally distributed continuous variables are presented as the mean ± standard deviation and were analyzed using Student’s t test. The Mann-Whitney U test was used for non-normally distributed continuous variables. Categorical variables are expressed as a frequency (percentage) and were analyzed using the Pearson chisquare test. The Wilcoxon Signed-Rank test was used to compare continuous variables. A p value < 0.05 was considered to indicate statistical significance.

### Data Measurement and Acquisition

Demographic data including age, height, weight, and American Society of Anesthesiologists (ASA) rating were recorded for all cases. The main endpoints of the study were the patient’s recovery time from anesthesia (the time required for the subject’s MOAA/S(Chernik et al. 1990) score to be 5 points since stopping the use of remimazolam or propofol) and the time to hospital discharge (the time required for the subject’s PADS(Palumbo et al. 2013) score to express 9 points since stopping the use of remimazolam or propofol). The secondary endpoints were the success of the first dose of sedation (1 minute after the administration of the initial dose of the drug, the eyelashes of the patient were stimulated with a cotton swab, and the sedation was considered successful if there was no reflex), the occurrence of hypotension (the blood pressure during anesthesia decreased by more than 20% compared with the systolic blood pressure during the fully awake period or the systolic blood pressure decreased by more than 20%. If the systolic blood pressure drops to ≤80mmHg, hypotension is considered to be present) and injection pain occurs (when the initial dose of drug is given, involuntary retraction of the limb on the injection side or the patient complains of pain in the limb on the injection side is considered to be pain).

## Results

From July 28, 2021 to September 2, 2021, a total of 155 patients were included in this study, and 14 cases were excluded due to reasons such as high or low BMI, anemia, patients receiving assisted reproductive technology or breastfeeding. 46 cases were excluded and withdrawn due to patients or family members refusing to participate in the study or incomplete relevant information. A total of 95 patients were finally analyzed in this study (Fig.1).

**Figure 1:**
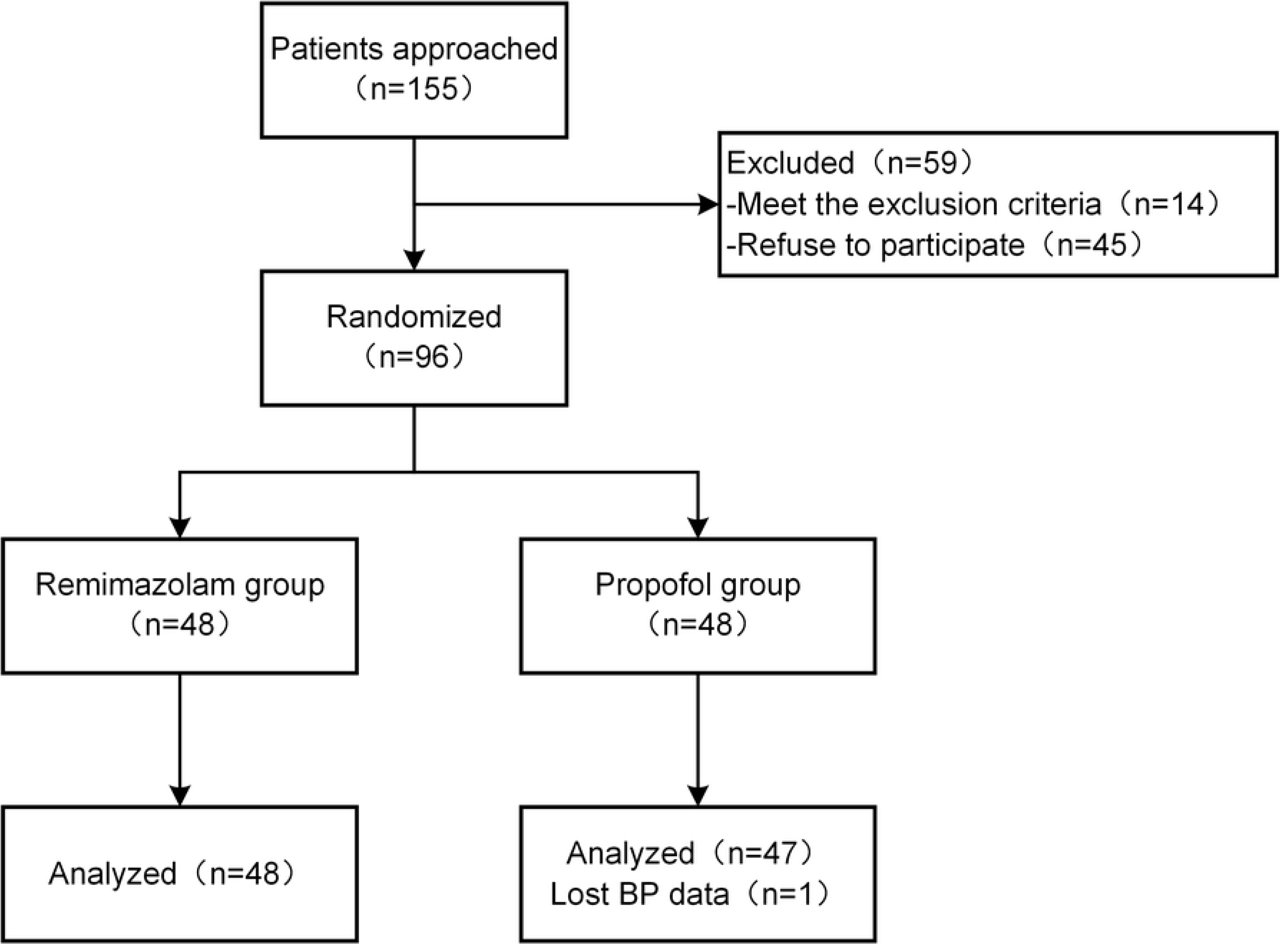
Flow Diagram.

Demographic data are detailed in Table 1, age (*P =*0.825), height (*P =*0.174), weight(*P =*0.667) and ASA grade(*P* =0.972) were not significantly different between the two groups.

**Table 1:**
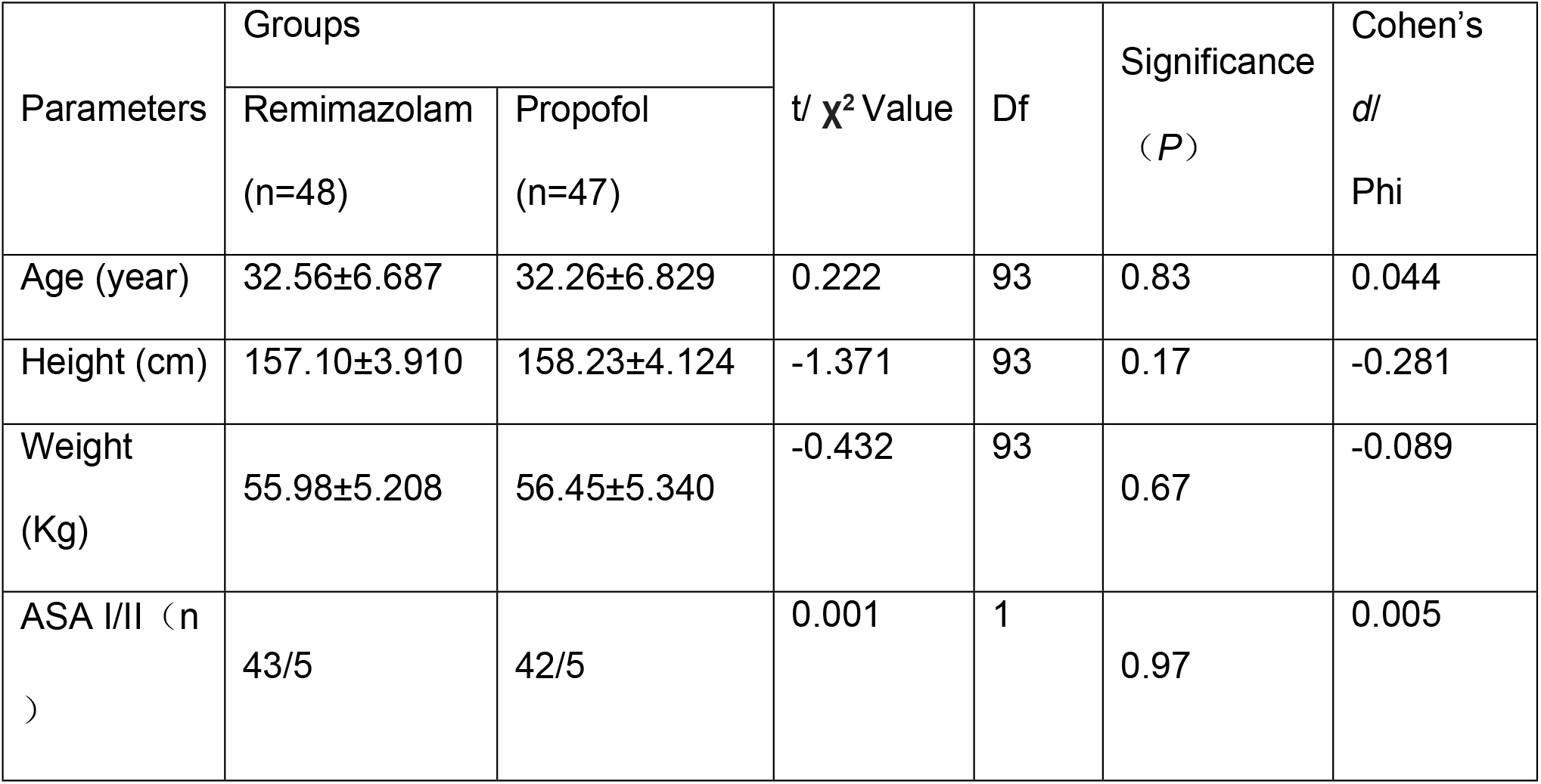
Demographic data (N = 95). All data are expressed as means ± SD.

| Parameters | Groups | | t/ $\chi^2$ Value | Df | Significance<br>($P$) | Cohen's<br>d/<br>Phi |
| --- | --- | --- | --- | --- | --- | --- |
|  | Remimazolam<br>(n=48) | Propofol<br>(n=47) |  |  |  |  |
| Age (year) | 32.56±6.687 | 32.26±6.829 | 0.222 | 93 | 0.83 | 0.044 |
| Height (cm) | 157.10±3.910 | 158.23±4.124 | -1.371 | 93 | 0.17 | -0.281 |
| Weight<br>(Kg) | 55.98±5.208 | 56.45±5.340 | -0.432 | 93 | 0.67 | -0.089 |
| ASA I/II (n<br>) | 43/5 | 42/5 | 0.001 | 1 | 0.97 | 0.005 |

As for the primary endpoint, the recovery time from anesthesia in group R was significantly shorter than that in group P (*P =*0.019), and the data kurtosis in group R was higher than that in group P(1.19 vs −0.56), but there was no significant difference in the time to hospital discharge (*P =*0.696).In secondary endpoints, there was no significant difference in the success rate of the first dose of sedation between the two groups (*P =*0.362), but the incidence of hypotension during anesthesia (*P=*0.001) and the incidence of injection pain (*P =*0.001) in group R were significantly lower than those in group R. (Table 2)

**Table 2:** Measurement data (N = 95). All data are expressed as means ± SD.

| Parameters | Groups | | t/ $\chi^2$<br>Value | Df | Significance<br>($P$) | Cohen's d/<br>Phi |
| --- | --- | --- | --- | --- | --- | --- |
|  | Remimazolam<br>(n=48) | Propofol<br>(n=47) |  |  |  |  |
| Anesthesia recovery time(min) | 6.23±1.547 | 7.09±1.932 | -2.386 | 93 | 0.019 | -0.491 |
| Discharge time(min) | 89.58±18.669 | 91.23±22.235 | -0.392 | 93 | 0.696 | -0.804 |
| First dose sedation success(n) | 44(91.7%) | 46(97.9%) | 1.834 | 1 | 0.362 | 0.601 |
| Hypotension(n) | 10(20.8%) | 26(55.3%) | 12 | 1 | 0.001 | 3.364 |
| Injection pain(n) | 6(12.5%) | 32(68.1%) | 30.572 | 1 | 0.001 | 5.529 |

## Discussion

Through this study, it can be found that remimazolam tosilate is used as the initial dose of 0.2 mg/kg in hysteroscopy anesthesia, which has a sedative success rate not inferior to that of propofol, and the average recovery time is shorter and more concentrated. Suggesting that when it is used for hysteroscopic anesthesia, patients need to stay in the PACU for a shorter time, and the drug metabolism is more predictable, which is beneficial to reduce the burden on PACU doctors and nurses and improve turnaround efficiency. The lower incidence of intraoperative hypotension in patients taking concomitant remimazolam tosilate may be a potential benefit of remimazolam, because intraoperative hypotension caused by anesthetic drugs has been shown to be associated with various postoperative organ dysfunction and adverse outcomes(Wesselink et al. 2018). In addition, the less incidence of injection pain may give remimazolam tosilate an advantage in special patients (eg, children)(Nyman et al. 2005). This study did not find that remimazolam tosilate was associated with the shortening of hospital discharge time, which may be due to the limitations of the design of this study, for example, this study was a single-center study with a small sample size, and no propensity scoring or weighted statistics were performed on patients’ preoperative fasting time or individual nutritional status.

## Conclusions

Remimazolam tosilate is suitable for hysteroscopy anesthesia.

Remimazolam tosilate can reduce intraoperative hypotension and injection pain during examination sedation.

Recovery time may be further shortened with specific antagonist flumazenil.

More optimal dosage or target-controlled infusion model still needs to be further explored.

## Data Availability

All relevant data are within the manuscript and its Supporting Information files.

## Acknowledgements

The author thanks the anesthetist nurses for their dedication to patient observation and data collection

## Declarations

The data that supports the findings of this study are available within supplementary material. No potential competing interest was reported by the authors.

The authors received no financial support for the research, authorship, or publication of this article.

Xiangyi Lin and Shengjie Chen wrote the main manuscript text, Huiyan Liang collected and counted all the clinical data, all authors reviewed the manuscript.

